# Long-term neurological manifestations of COVID-19: prevalence and predictive factors

**DOI:** 10.1101/2020.12.27.20248903

**Authors:** Andrea Pilotto, Viviana Cristillo, Stefano Cotti Piccinelli, Nicola Zoppi, Giulio Bonzi, Davide Sattin, Silvia Schiavolin, Alberto Raggi, Antonio Canale, Stefano Gipponi, Ilenia Libri, Martina Frigerio, Michela Bezzi, Matilde Leonardi, Alessandro Padovani

## Abstract

**Background:** Clinical investigations have argued for long-term neurological manifestations in both hospitalized and non-hospitalized COVID-19 patients. It is unclear whether long-term neurological symptoms and features depend on COVID-19 severity.

**Methods:** from a sample of 208 consecutive non-neurological patients hospitalized for COVID-19 disease, 165 survivors were re-assessed at 6 months according to a structured standardized clinical protocol. Prevalence and predictors of long-term neurological manifestations were evaluated using multivariate logistic regression analyses.

**Results:** At 6-month follow-up after hospitalisation due to COVID-19 disease, patients displayed a wide array of symptoms; fatigue (34%), memory/attention (31%), and sleep disorders (30%) were the most frequent. At neurological examination, 40% of patients exhibited neurological abnormalities, such as hyposmia (18.0%), cognitive deficits (17.5%), postural tremor (13.8%) and subtle motor/sensory deficits (7.6%). Older age, premorbid comorbidities and severity of COVID-19 were independent predictors of neurological manifestations in logistic regression analyses.

**Conclusions:** premorbid vulnerability and severity of SARS-CoV-2 infection impact on prevalence and severity of long-term neurological manifestations.

## INTRODUCTION

After the first cases of the novel coronavirus disease 2019 (COVID-19) were reported in Wuhan, China, in December 2019, the spread rapidly in Europe became a pandemic, involving millions of cases worldwide^1^. With the increasing number of confirmed cases and the accumulating clinical data, it is now well established that, in addition to the predominant respiratory symptoms, a significant proportion of COVID-19 patients experience neurological symptoms and syndromes^2-5^. Clinical findings on previously hospitalized and non-hospitalized patients with COVID-19 reported the persistence of multiple symptoms, particularly fatigue, dyspnoea, sleep disturbances and memory complaints^6-8^. Accordingly, some authors have suggested the so called, but not yet defined, “post-COVID-19 syndrome” based on persistent symptoms reported after resolution of SARS-CoV-2 infection^8^. To date, however, the prevalence and severity of neurological long-term manifestations and the correlation with severity of COVID-19 infection are still under debate, as no long-term data using extensive neurological assessment are still available yet in the growing literature.

In this study, subjects previously hospitalised for COVID-19 disease entered a longitudinal study in order to evaluate general and neurological manifestations after 6 months of follow-up and their potential relationship with pre-morbid conditions and severity of respiratory infection.

## METHODS

All patients who survived COVID-19 disease and were discharged between February and April 2020 from a COVID-19 Unit of the ASST Spedali Civili Brescia Hospital were asked to participate to a follow-up study. This includes a standardised evaluation of medical history, self-reported neurological symptoms and a complete neurological examination at 6 months. Premorbid conditions were recorded at admission using the Cumulative Illness rating scale^9^ (CIRS). Patients with premorbid neurological conditions (including dementia) were excluded. Hospitalisation data included the severity of COVID-19 disease, classified according to the Brescia-COVID Respiratory Severity Scale (BCRSS), stratifying patients into mild, moderate and severe^10^ and the quick Sequential Organ Failure Assessment (qSOFA) score^11^.

At follow-up, data were collected using a checklist of neurological symptoms related to central, peripheral, myopathic and cognitive manifestations, whereas the neurological examination assessed cranial nerves, motor (global and focal), sensory, cerebellar, basal ganglia-related function, deep tendon reflexes, pyramidal signs. Global cognitive function was carried out by using the Montreal Cognitive Assessment (MoCA) by Italian validated norms^12^.

The study was approved by the local ethics committee of ASST “Spedali Civili di Brescia” Hospital and the requirement for informed consent was waived by the Ethics Commission (NP 4166).

### Statistical analysis

Differences between patients according to COVID-19 respiratory severity (BCRSS) and the association with neurological complaints were evaluated by Fisher’s exact test or ANOVA with Bonferroni correction for dichotomic and continuous variables, respectively. To explore the risk factors associated with neurological symptoms and neurological features, univariable and multivariable logistic regression models were implemented, including the following predictors: age, sex, premorbid CIRS, days of hospitalisation and COVID-19 severity.

The data that support the findings of this study are available from the corresponding author upon reasonable request.

## RESULTS

From a sample of 208 consecutively hospitalized patients for COVID-19 disease, 33 deceased during hospitalisation. Survivors were younger (p=0.001; 65.7±12.6 vs 78.6±8.6) and exhibited less comorbidities (p=0.004, mean CIRS 1.36±0.51 vs 1.51±0.4) and lower COVID-19 severity (p=0.001, mean BCRSS 0.90±0.85 vs 2.89 ± 0.32) compared to deceased patients.

Out of 175 survivors, five patients died after discharge, three had a previous diagnosis of dementia and two refused to participate, resulting in a final sample of 165 patients (supplementary Figure 1). Patients stratified according to COVID-19 severity (BCRSS) differ for number of days of hospitalisation, O2 treatment and qSOFA but not for age or premorbid CIRS (Table 1).

**Table 1.** Demographic and clinical characteristics of the sample according to COVID-19 severity.

|  | **Total**<br>**(n=165)** | **Mild**<br>**(n=57)** | **Moderate**<br>**(n=77)** | **Severe**<br>**(n=31)** | ***p* value** |
| --- | --- | --- | --- | --- | --- |
| Age, years | 64.8 ± 12.6 | 62.3 ± 13.1 | 67.3 ± 12.3 | 63.2 ± 11.2 | 0.06 |
| Sex, female n (%) | 50 (30.3%) | 22 (38.5%) | 18 (23.4%) | 10 (32.2%) | 0.16 |
| Days of Hospitalization | 11.6 ± 8.8 | 7.8 ± 4.2 | 12.3 ± 9.4 | 17.1 ± 10.4 | **0.001#¶\*** |
| Oxygen therapy, n (%) | 128 (77.6%) | 20 (35.1%) | 77 (100%) | 31 (100%) | **0.001¶** |
| Non-invasive ventilation, n (%) | 18 (11.0%) | 0 (0%) | 0 (0%) | 18 (58.3%) | **0.001#¶** |
| Intubation, n (%) | 2 (1.2%) | 0 (0%) | 0 (0%) | 2 (6.4%) | **0.001#¶** |
| qSOFA | 0.44 ± 0.53 | 0.0 ± 0.0 | 0.48 ± 0.50 | 1.10 ± 0.29 | **0.001#¶\*** |
| CIRS pre total | 18.6 ± 3.3 | 18.1 ± 3.2 | 19.4 ± 3.6 | 17.9 ± 2.8 | 0.03 |
| CIRS, pre severity mean | 1.35 ± 0.25 | 1.31 ± 0.24 | 1.40 ± 0.26 | 1.28 ± 0.20 | 0.02 |

At follow-up, no patients reported new hospitalisations or new-onset of medical conditions. The most common symptoms reported were fatigue (34%), memory complaints (31%), sleep disorders (30.8%) and myalgias (29.6%), followed by depression/anxiety, visual disturbances, paraesthesia and hyposmia (Figure 1, supplementary Table 1). Patients with moderate/severe COVID-19 reported higher number of symptoms at follow-up (p=0.004) after correction for age and premorbid CIRS. In univariable analyses, moderate/severe COVID-19 was associated with increased risk of memory complaints (OR 2.6, 95% CI 1.18-5.8), loss of dependency in IADL (OR 2.6, 95% CI 1.12-6.2), confusion (OR 2.9, 95% CI 1.12-7.8), fatigue (OR 2.1, 95% CI 0.95-4.6), and visual disturbances (OR 3.5, 95% CI 1.5-8.4) at 6-months of follow-up compared to mild disease.

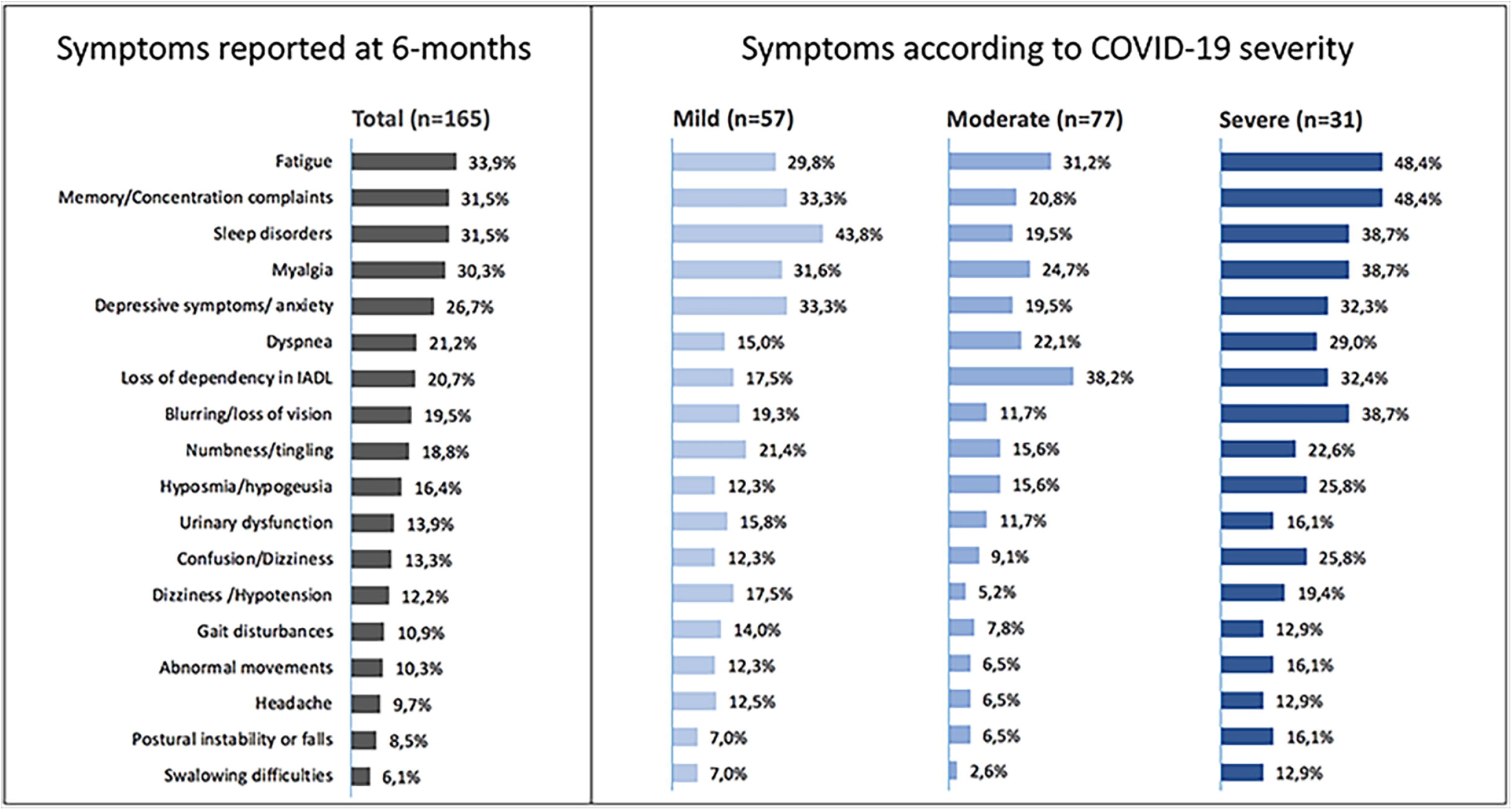

Multivariable analyses identified premorbid comorbidities (p=0.006), age at admission (p=0.048) and severity of COVID-19 (p=0.04) as predictors of total number of symptoms reported.

One-hundred and five patients (63,6%) were further evaluated using a standard neurological examination and cognitive screening. This group of patients were comparable to patients evaluated via telemedicine for age, sex distribution, pre-morbid CIRS, severity of COVID-19 and reported neurological symptom. At standard examination, 42/105 (40%) exhibited neurological abnormalities, including subjective dysgeusia/hyposmia (n=19), enhanced physiological tremor (n=15) and cognitive impairment (n=17). Ten patients exhibited low-limb reduced deep tendon reflexes, two of them exhibited isolated feet hypoesthesia and two reached a diagnosis of sensor-motor polyneuropathy with distal mild sensory and motor deficits (bilateral deficits within the extension of feet 4/5 MRC). No alterations within cranial nerves, cerebellar function or pyramidal signs were detected.

Neurological abnormalities at examination were associated with older age (p=0.005), higher number of premorbid comorbidities (p=0.001), higher COVID-19 severity (p=0.05), longer hospitalisation (p=0.002) and higher number of neurological symptoms reported (p=0.007). Logistic regression identified duration of hospitalisation (p=0.02) and premorbid comorbidities(p=0.03) as the best predictors of neurological abnormalities at examination.

## DISCUSSION

Findings showed that previously hospitalized COVID-19 patients reported a wide array of neurological symptoms six months after SARS-CoV-2 infection, predicted by combination of age, premorbid conditions and severity of disease.

These data extend recent studies which have argued for a high prevalence of post-COVID clinical manifestations and claimed that long-term consequences of COVID-19 involve both central and peripheral nervous systems^6-8,13,14^.

In the present cohort of patients with six months of follow-up and extensive neurological evaluation, the most prevalent symptoms reported were fatigue, memory complaints, sleep disorders and myalgias followed by depression/anxiety, visual disturbances, paraesthesia and hyposmia. These findings extended the earlier works of Carfi^7^ and Goertz^8^, reporting a high persistence of respiratory symptoms and fatigue in more than half of hospitalized and non-hospitalized patients three months after COVID-19 infection. In our study, long-term neurological complaints showed different distributions according to COVID-19 severity, which was associated with increased total number of symptoms, fatigue, memory complaints and confusion. The impact of SARS-CoV-2 severity in long-term persistent respiratory symptoms and fatigue has been recently highlighted by a large survey 6 months after discharge in Wuhan^6^. Our study, conducted in smaller but more homogeneous older population consecutively enrolled in a single COVID-19 unit, suggest that even small differences in the severity of infection can impact on long-term neurological manifestations. This effect was prominent for cognitive symptoms, including memory complaints and attention deficits-whereas other symptoms- - such as myalgias or numbness - appeared to be independent from severity. This might suggest a higher vulnerability of central nervous system to severe SARS-CoV-2 infection possibly through higher general inflammatory response and longer hospitalisation- as already highlighted for other infectious disease^16^. We found age and premorbid comorbidities as important independent predictors of long-term symptoms, at variance with the data reported by Huang and coauthors for respiratory symptoms and fatigue^6^. These findings are of particular interest, as we excluded patients with premorbid dementia or other known neurological conditions or patients with neurologic COVID-19 presentation^5^, focusing on neurological features in patients with prominent respiratory COVID-19 disease^16^.

At standardised neurological examination, 40% of subjects exhibited mild abnormalities -not reported before the hospitalisation for COVID-19 disease. The most prevalent features were hyposmia, cognitive impairment, enhanced physiological tremor and subtle motor and sensory low-limb deficits in several cases. Age, premorbid comorbidities and longer hospitalization were, again, the strongest predictors of neurological abnormalities at examination. This would suggest that SARS-CoV-2 infection has probably a stronger long-term neurological impact in older subjects with higher vulnerability who suffered an acute respiratory syndrome. The complex interaction between physical and brain resilience and long-term disability after hospitalization has been indeed already demonstrated for other infectious diseases, such as community-acquired pneumonia^16^. In fact, we need to acknowledge that the experience of hospitalization due to COVID-19 infection in frail elderly should is an important confounding factor-as it can influence the relationship between long-term manifestations and psychosocial long-term disturbances^17-19^.

Several limitations should be acknowledged. First, pre-morbid conditions were based on medical records and assessment during hospitalisation thus not allowing a complete premorbid neurological screening. Second, we excluded from the study patients with neurological disorders before or concomitant the acute phase of SARS-CoV-2 infection, thus potentially underestimate the global neurological burden due to COVID-19. Furthermore, this is a single-centre study conducted in an homogeneous cohort of patients with moderate severity of COVID-19 and large studies including intubated and non-hospitalized patients are warranted to confirm these findings.

Limitations notwithstanding, our findings indicate that several neurological features are a relevant component of long-term manifestations of COVID-19 disease especially in more vulnerable and severe patients, thus underlying the clinical need for longitudinal programs able to track the real impact of SARS-CoV-2 infection on long-term brain health status^18-21,^.

## ACKNOWLEDGEMENT

The authors thanks all the participant for their participation.

---

**Supplementary Table 1:** Different prevalence of Neurological symptoms reported by patients using the checklist according to different COVID-19 severity. Abbreviations: BCRSS, Brescia-COVID Respiratory Severity Scale; IADL, instrumental activities of daily living

|  | **Total**<br>**(n=165)** | **Mild**<br>**(n=57)** | **Moderate**<br>**(n=77)** | **Severe**<br>**(n=31)** | ***p* value** |
| --- | --- | --- | --- | --- | --- |
| Fatigue | 56 (33.9%) | 17 (29.8%) | 24 (31.2%) | 15 (48.4%) | 0.18 |
| Memory/Concentration complaints | 52 (31.5%) | 19 (33.3%) | 16 (20.8%) | 15 (48.4%) | 0.015 |
| Sleep disorders | 52 (31.5%) | 25 (43.8%) | 15 (19.5%) | 12 (38.7%) | 0.006 |
| Myalgia | 50 (30.3%) | 18 (31.6%) | 19 (24.7%) | 12 (38.7%) | 0.319 |
| Depressive symptoms/ anxiety | 44 (26.7%) | 19 (33.3%) | 15 (19.5%) | 10 (32.3%) | 0.134 |
| Dyspnea | 35 (21.2%) | 9 (15.7%) | 17 (22.1%) | 9 (29.0%) | 0.28 |
| Loss of dependency in IADL | 34 (20.7%) | 10 (17.5%) | 13 (16.8%) | 11 (32.4%) | 0.08 |
| Blurring/loss of vision | 32 (19.5%) | 11 (19.3%) | 9 (11.7%) | 12 (38.7%) | 0.006 |
| Numbness/tingling | 31 (18.8%) | 12 (21.4%) | 12 (15.6%) | 7 (22.6%) | 0.58 |
| Hyposmia/hypogeusia | 27 (16.4%) | 7 (12.3%) | 12 (15.6%) | 8 (25.8%) | 0.26 |
| Urinary dysfunction | 23 (13.9%) | 9 (15.8%) | 9 (11.7%) | 5 (16.1%) | 0.72 |
| Confusion/Dizziness | 22 (13.3%) | 7 (12.3%) | 7 (9.1%) | 8 (25.8%) | 0.07 |
| Dizziness /Hypotension | 20 (12.2%) | 10 (17.5%) | 4 (5.2%) | 6 (19.4%) | 0.04 |
| Gait disturbances | 18 (10.9%) | 8 (14.0%) | 6 (7.8%) | 4 (12.9%) | 0.46 |
| Abnormal movements | 17 (10.3%) | 7 (12.3%) | 5 (6.5%) | 5 (16.1%) | 0.67 |
| Headache | 16 (9.7%) | 7 (12.5%) | 5 (6.5%) | 4 (12.9%) | 0.41 |
| Postural instability or falls | 14 (8.5%) | 4 (7.0%) | 5 (6.5%) | 5 (16.1%) | 0.24 |
| Swallowing difficulties | 10 (6.1%) | 4 (7.0%) | 2 (2.6%) | 4 (12.9%) | 0.11 |

p values were calculated by chi-square test.

