## Supplementary figures and images for "Long-term neurological manifestations of COVID-19: prevalence and predictive factors"

### supplementary figure 1

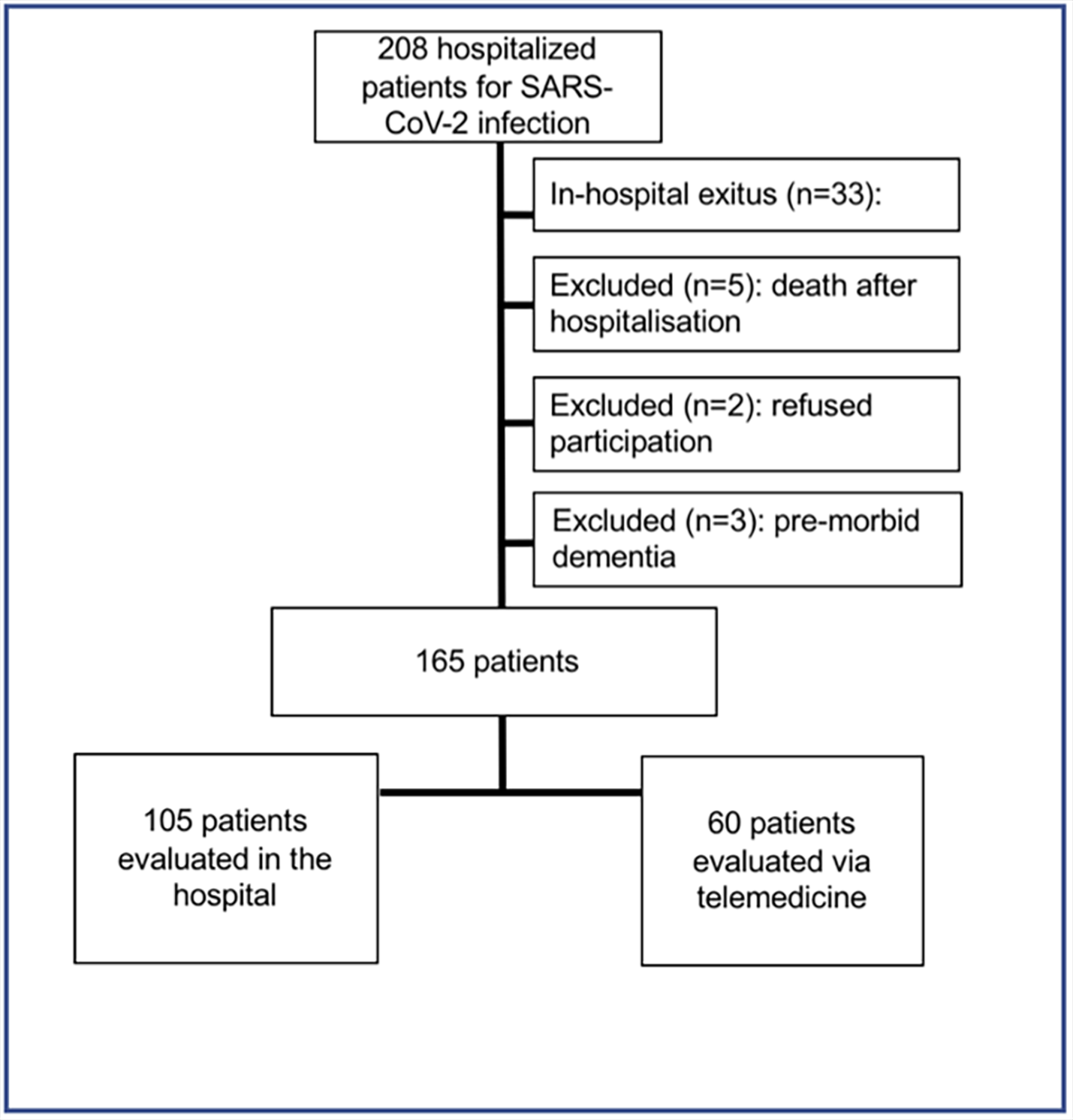
